# Improving Breast Cancer Detection in Higher Risk Women: A Multimodality Imaging Evaluation in a Private Screening Clinic

**DOI:** 10.1101/2025.09.08.25334122

**Authors:** Sugania Reddy, MERCY RADIOLOGY and Breast Clinic (ALLEVIA Radiology and Breast Institute) AND PILOT TEAM, Nicholas Knowlton, Annette Lasham

## Abstract

**Introduction:** While 2D mammography is the standard for breast cancer screening, its sensitivity is reduced in dense breasts, impacting early detection and extent assessment. This study, for the first time in New Zealand, evaluated the utility of supplementary multi-modality imaging (tomosynthesis, ultrasound, and MRI) in a risk-stratified population.

**Methods:** A retrospective case study (May 2022 – September 2023, New Zealand private clinic) analysed patients by Tyrer-Cuzick (TC) v8 lifetime risk and by Volpara density categories. All patients in the screening pathway (n = 2171) underwent 2D mammography and tomosynthesis. Those patients with high breast density received supplementary ultrasound, and those with TC8 risk scores of ≥ 30% were offered abbreviated MRI. Symptomatic patients (n = 230) underwent standard diagnostic workup. Detection rates and extent of disease using multimodality imaging were compared.

**Results:** Of the 2401 patients, 205 were high-risk criteria (≥30%) and 19 breast cancers (16 invasive, 3 DCIS only) were diagnosed. Tomosynthesis identified 38% (6/16) more invasive cancers than 2D mammography alone. Ultrasound and MRI detected an additional 27% (4/16) invasive cancers occult on other modalities, predominantly in those women with density D breasts. Ultrasound and particularly MRI demonstrated superior accuracy in assessing disease extent, including identifying multifocal and multicentric disease that was not detected by 2D or Tomosynthesis.

**Conclusion:** Supplementary screening modalities, particularly MRI, significantly improve breast cancer detection and assessment of disease extent in high-risk women. These findings support a personalized screening approach integrating risk assessment and breast density to guide imaging modality selection.

## Introduction

Breast cancer remains a leading cause of cancer-related morbidity and mortality in women globally. ^1^ A recent study found New Zealand has one of the highest incidences of breast cancer worldwide. ^2^ Population-based screening with 2D digital mammography has reduced mortality, but diagnostic sensitivity is limited, especially in women with dense fibroglandular stroma. ^3^ Dense breast tissue both elevates cancer risk and obscures lesions on mammography, increasing the chance of missed or underestimated disease. ^4, 5^

To address this, supplementary imaging modalities have been developed and are increasingly integrated into clinical care. Digital breast tomosynthesis generates quasi-3D reconstructions from multiple low-dose projections, improving lesion conspicuity. ^6^ Whole-breast ultrasound is radiation-free and enhances cancer detection in dense breasts. ^7^ Breast magnetic resonance imaging (MRI) provides the highest sensitivity, detecting subtle lesions and defining disease extent via contrast enhancement. ^8^

Risk stratification has advanced personalised screening withmodels like Tyrer-Cuzick v8 (TC8), which estimates lifetime breast cancer risk from family history, hormonal/reproductive factors, and density. ^9, 10^ These models help determine eligibility for adjunctive imaging beyond mammography.

However, limited evidence exists on how risk and breast density interact to affect detection yield, especially in New Zealand women. ^11, 12^ This study evaluated tomosynthesis, ultrasound, and abbreviated breast MRI in a real-world screening population, stratified by risk and breast density. We hypothesised that supplementary modalities would enhance detection, especially in dense breasts or elevated-risk women, and better delineate disease extent compared to mammography alone.

## Methods

This was a retrospective cohort study conducted at Mercy Breast Clinic, a private multidisciplinary breast imaging centre in Auckland, New Zealand. The clinic provides a personalised breast screening and diagnostic service to asymptomatic and symptomatic women, outside of the national breast screening programme. Data were collected from consecutive women with Southern Cross Health insurance who underwent breast imaging between May 2022 and September 2023. All women gave their informed consent to participate in this study. Analysis of the study data was approved by AHREC (#27480).

Women were included if they had completed 2D digital mammography and tomosynthesis, with or without additional ultrasound or MRI as part of either a routine screening or diagnostic pathway. Women with a personal history of breast cancer were excluded. The final cohort included 2,401 women: 2,170 from the screening pathway and 230 from the diagnostic (symptomatic) pathway. A description of patient inclusion, pathway assignment, and imaging modality use is shown in Figure 1.

**Figure 1.**
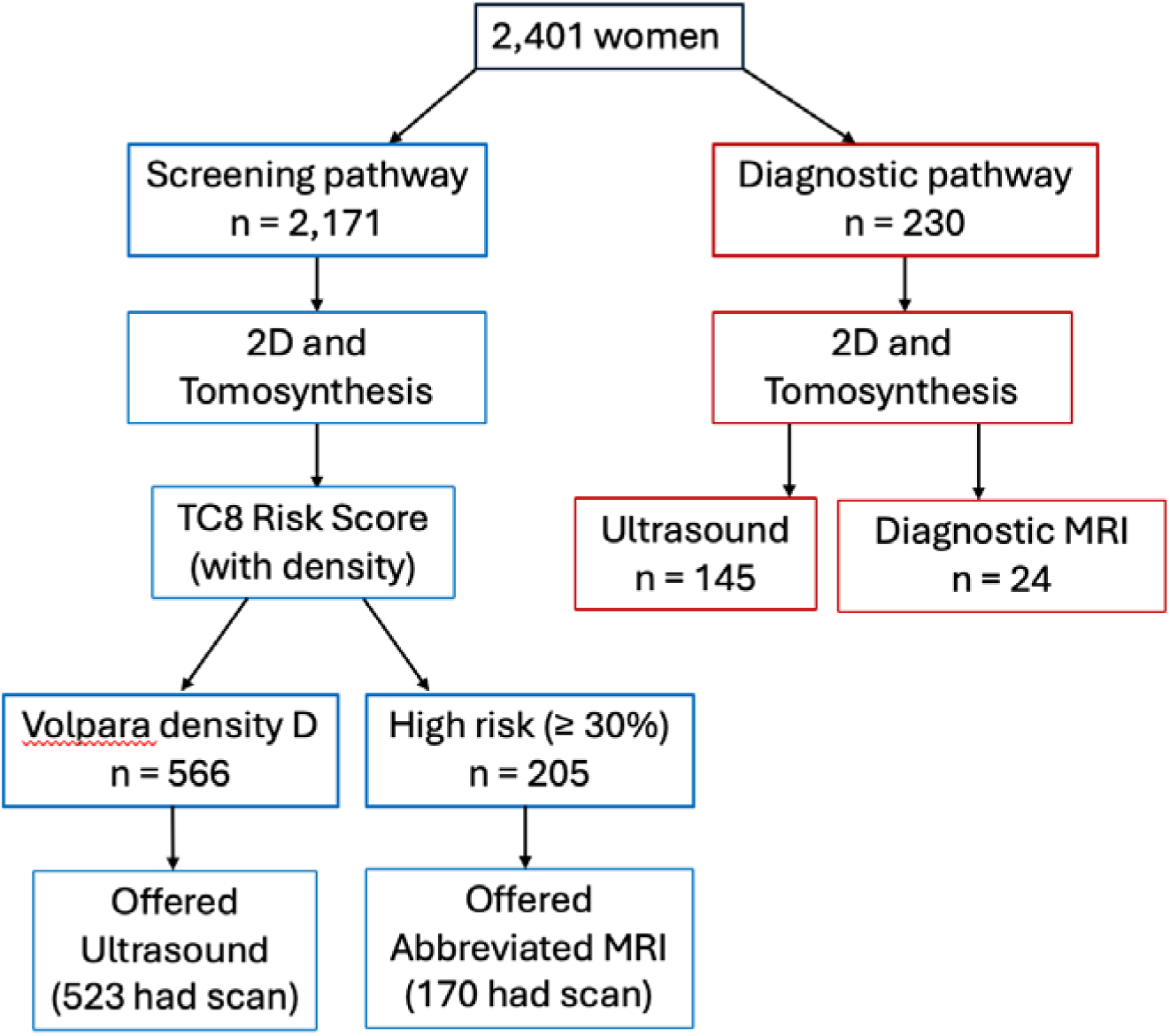
Flowchart of study cohort, imaging pathways, and breast cancer detection. A total of 2,401 women underwent breast imaging between May 2022 and September 2023. Patients were triaged to either the screening pathway (blue) or diagnostic pathway (red). All women received 2D mammography and tomosynthesis. Women with extremely dense breasts (Volpara density group D) were offered supplementary ultrasound. High-risk women (as defined by TC8 risk score of ≥ 30%) were eligible for abbreviated breast MRI. Cancers detected were stratified by invasive or *in situ* histology and mapped to the imaging modality on which they were first detected. The flowchart summarises pathway allocation and cancer detection by modality.

In the screening pathway, all women underwent bilateral 2D full-field digital mammography and tomosynthesis using Hologic and Siemens systems. Volumetric breast density was measured using Volpara® software, and women were assigned to a breast density group from A to D, based on predefined cut-offs and BIRADS breast density classification. ^13^ Women with extremely dense breasts (Group D) were offered whole-breast ultrasound, performed with Philips EPIQ systems using high-frequency linear transducers.

Women classified as “high risk” (defined below) were offered a novel abbreviated breast MRI protocol on a Siemens 3T scanner using Gadovist contrast (0.1 mL/kg, Bayer). This protocol represented a hybrid model, combining elements from both abbreviated MRI and ultrafast or FAST MRI techniques. ^14, 15^ The imaging sequences included axial T2-weighted, axial T1-weighted non–fat saturated, a dynamic TWIST (Time-resolved angiography With Interleaved Stochastic Trajectories) post-contrast T1-weighted fat-saturated series, a delayed 90-second axial T1 fat-saturated scan, and maximum intensity projection (MIP) images.

This combined protocol achieved a markedly reduced scan time of approximately 10 minutes, compared to the standard 40-minute protocol, while maintaining high sensitivity for breast cancer detection. Ultrafast acquisition via TWIST enabled lesion enhancement to be assessed immediately following thoracic aortic enhancement and prior to background parenchymal enhancement, which typically masks small or subtle lesions. This early imaging window enhances lesion conspicuity and improves spatial delineation, which is especially important in dense breasts, and has been shown to increase both specificity and reader efficiency. ^14, 15^ The protocol used in this study was adapted from ^14^, with the key modification of omitting diffusion-weighted imaging (DWI). This omission, based on limited diagnostic yield, enabled further reduction in scan time while preserving diagnostic performance.

Risk assessment was performed using the Tyrer-Cuzick version 8 (TC8) assessment, which estimates lifetime breast cancer risk based on family history, hormonal/reproductive factors, and breast density. ^10^ Note: the definition of family history in our study is having a first degree relative with either breast or ovarian cancer. Women have their lifetime risk classified as Average if their score is < 15%, Intermediate for those scoring >= 15 to < 20% and High lifetime risk if their score is ≥ 20%. ^16^To align our analysis with the National Institute for Health and Care Excellence (NICE) criteria commonly applied in Australia and New Zealand, we also evaluated an alternative stratification: Average (<15%), Intermediate (15–29.9%), and High (≥30%). In this manuscript we refer to these thresholds as the “TC8 Risk ANZ categories” to distinguish them from the original TC8 cut-offs while acknowledging that they are directly derived from NICE criteria. ^17^

The diagnostic pathway applied to women referred with symptoms or abnormal findings. These women received tailored imaging depending on presentation (Figure 1), but most underwent 2D mammography, tomosynthesis, and targeted ultrasound. MRI was selectively performed in cases of diagnostic uncertainty, suspected multifocal disease, or pre-operative staging.

For each patient, breast cancer diagnosis (invasive or *in situ* disease) and detection modality were recorded. Where cancers were visible on more than one modality, the first modality on which the cancer was visible was recorded. In cases where multiple imaging modalities were used, radiology reports were reviewed to assess whether each additional modality contributed to increased assessment of disease extent (e.g., greater lesion size, multifocality, or contralateral disease).

Data were analysed descriptively. Breast cancer detection counts were stratified by modality and grouped by TC8 Risk (standard and New Zealand thresholds), and breast density. Where applicable, comparisons were made between modalities for their contribution to detection and disease extent. No formal statistical hypothesis testing was performed due to sample size limitations in some subgroups.

## Results

“Using the Tyrer-Cuzick v8 (TC8) model, we evaluated how many women in the screening pathway cohort were classified into each of the three risk groups (Table 1). Based on the standard TC8 definition (≥20% lifetime risk), 537 women (approximately one-quarter of the cohort) were classified as high risk. This proportion was considered too large to be clinically practical as a stratification threshold, as it would result in an excessive number of women being directed toward supplementary screening. To address this, we applied the NICE criteria, which use a higher threshold (≥30% lifetime risk) to define the high-risk category. We refer to these thresholds in this manuscript as the ‘TC8 Risk ANZ categories’ to distinguish them from the original TC8 definitions.”

**Table 1.** Comparison of TC8 Risk (Tyler Cuzick v8) and TC8 Risk ANZ Categories.

| **TC8 Risk Category** | **Average**<br>**(n=1318)** | **Intermediate**<br>**(n=316)** | **High Risk**<br>**(n=537)** |
| --- | --- | --- | --- |
| **TC8 Risk ANZ Category** |  |  |  |
| Average | 1318 (100%) | 0 (0%) | 0 (0%) |
| Intermediate | 0 (0%) | 316 (100%) | 332 (61.8%) |
| High | 0 (0%) | 0 (0%) | 205 (38.2%) |

Note: TC8 High Risk is $\geq 20\%$ lifetime risk, TC8 ANZ High Risk is $\geq 30\%$ lifetime risk

Next, we analysed basic demographics and breast cancer detection rates (invasive or *in situ*) across the TC8 Risk ANZ categories. Women classified as Intermediate or High Risk had significantly lower mean and median ages compared to those in the Average Risk category (Table 2). Most notably, breast density, as measured by both the Volpara Breast Density Score and Breast Density Group, was markedly higher in the Intermediate and especially the High Risk groups (Table 2).

**Table 2.** Analysis of screening pathway cohort by TC8 Risk ANZ Score.

| **TC8 Risk ANZ Category** | **Average**<br>**(n=1318)** | **Intermediate**<br>**(n=648)** | **High Risk**<br>**(n=205)** |
| --- | --- | --- | --- |
| **Age at Mammogram (years)** |  |  |  |
| Mean (SD) | 58.8 (10.5) | 50.5 (7.8) | 48.9 (6.2) |
| Median [Min, Max] | 59 [36, 83] | 49 [38, 79] | 49 [40, 78] |
| **Breast Density Score (Volpara)** |  |  |  |
| Mean (SD) | 7.83 (5.66) | 16.9 (10.1) | 22.3 (8.9) |
| Median [Min, Max] | 6.0 [1.8, 36.6] | 14.1 [2.0, 43.3] | 24.7 [2.6, 38.7] |
| **Breast Density Group** |  |  |  |
| A | 396 (30.0%) | 48 (7.4%) | 1 (0.5%) |
| B | 429 (32.5%) | 97 (15.0%) | 11 (5.4%) |
| C | 393 (29.8%) | 189 (29.2%) | 40 (19.5%) |
| D | 100 (7.6%) | 314 (48.5%) | 153 (74.6%) |
| **Cancer Detected?** |  |  |  |
| Yes | 8 (0.6%) | 4 (0.6%) | 2 (1.0%) |
| No | 1310 (99.4%) | 644 (99.4%) | 203 (99.0%) |

In the screening pathway cohort, a total of 14 breast cancers were detected (0.65%). Detection rates were slightly higher in the High Risk group, with 1% of women having a breast cancer detected, compared to 0.6% in the Average and Intermediate Risk group (Table 2).

### Breast Cancer Detection by Screening Modality

We analysed the screening pathway cohort to evaluate detection of invasive breast cancer by imaging modality, stratified by TC8 ANZ risk categories (Table 3). Given that the Average Risk was substantially larger than the Intermediate and High Risk groups, it is important to consider absolute numbers in context. 2D mammography alone detected four invasive cancers, all in women classified as TC8 ANZ “Average” risk. Tomosynthesis identified three additional cancers, which did not show typical features of malignancy on 2D mammography, highlighting this modality’s increased sensitivity of invasive cancer detection over conventional mammography (Table 3). Ultrasound detected an additional three cancers that were missed by 2D mammography and tomosynthesis, reinforcing its role as an adjunct screening tool.

**Table 3:**
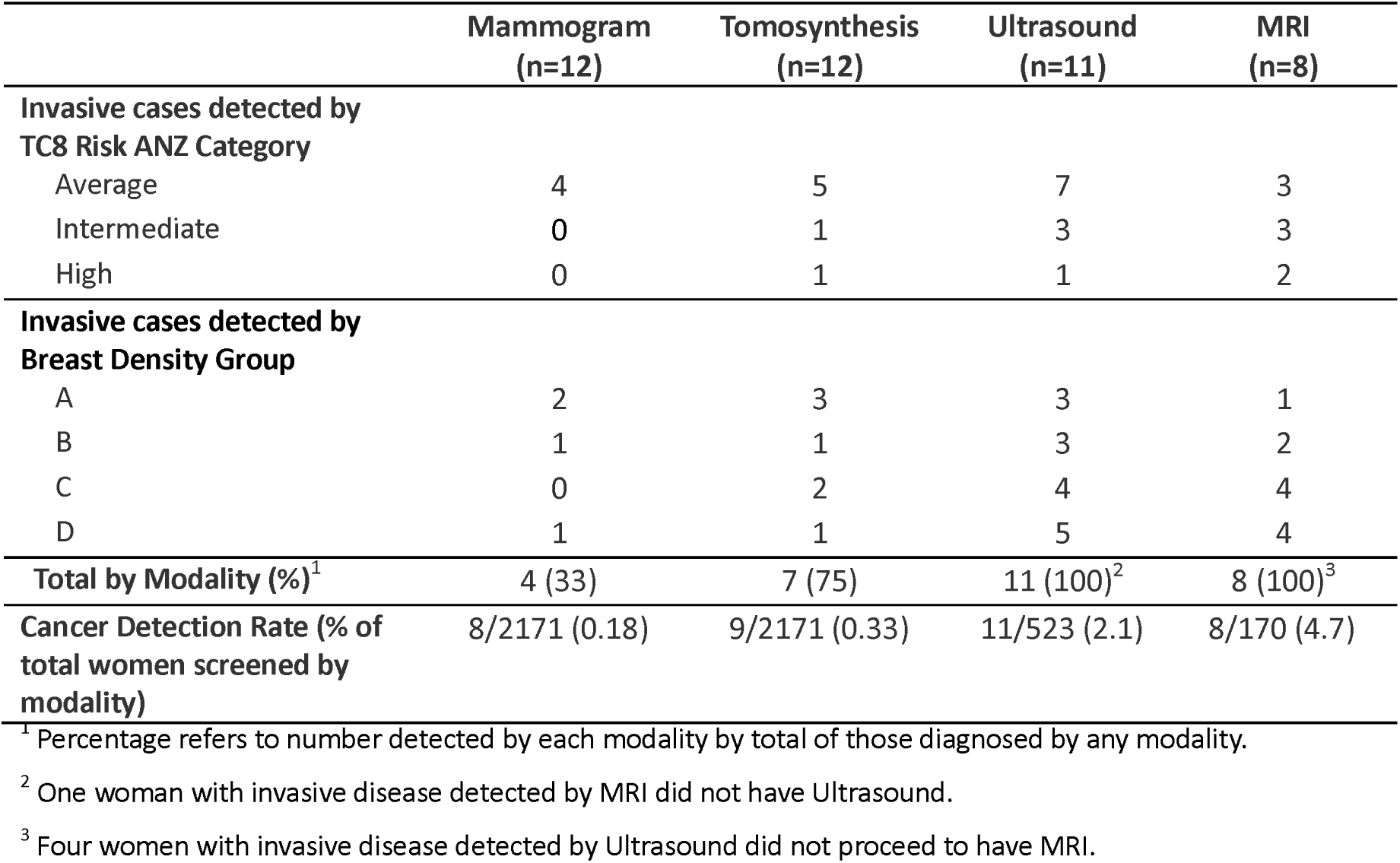
Analysis of the screening pathway cohort showing invasive breast cancer cases detected by each modality, by TC8 Risk ANZ Category or breast density.

|  | Mammogram<br>(n=12) | Tomosynthesis<br>(n=12) | Ultrasound<br>(n=11) | MRI<br>(n=8) |
| --- | --- | --- | --- | --- |
| **Invasive cases detected by TC8 Risk ANZ Category** |  |  |  |  |
| Average | 4 | 5 | 7 | 3 |
| Intermediate | 0 | 1 | 3 | 3 |
| High | 0 | 1 | 1 | 2 |
| **Invasive cases detected by Breast Density Group** |  |  |  |  |
| A | 2 | 3 | 3 | 1 |
| B | 1 | 1 | 3 | 2 |
| C | 0 | 2 | 4 | 4 |
| D | 1 | 1 | 5 | 4 |
| **Total by Modality (%)<sup>1</sup>** | 4 (33) | 7 (75) | 11 (100)<sup>2</sup> | 8 (100)<sup>3</sup> |
| **Cancer Detection Rate (% of total women screened by modality)** | 8/2171 (0.18) | 9/2171 (0.33) | 11/523 (2.1) | 8/170 (4.7) |

<sup>1</sup> Percentage refers to number detected by each modality by total of those diagnosed by any modality.
<sup>2</sup> One woman with invasive disease detected by MRI did not have Ultrasound.
<sup>3</sup> Four women with invasive disease detected by Ultrasound did not proceed to have MRI.

For women classified as High Risk (≥ 30% lifetime risk), we offered a novel abbreviated MRI protocol that combined elements of both FAST MRI and abbreviated MRI. This protocol incorporated ultrafast dynamic contrast-enhanced imaging, which captures early lesion enhancement immediately after thoracic aortic enhancement and prior to the onset of background parenchymal enhancement. This timing reduces the potential masking effect of background enhancement, which can otherwise obscure small or subtle lesions. As a result, this adapted protocol preserved the high sensitivity of MRI while improving lesion conspicuity and spatial delineation. It proved particularly effective for the eight women where invasive cancers were detected. MRI would likely have detected additional cases that were identified by ultrasound, but four women with invasive breast cancer did not undergo MRI following ultrasound screening. Together these findings suggest that the TC8 Risk ANZ categories did not strongly differentiate which women required additional screening beyond standard mammography to detect invasive breast cancer.

The screening pathway cohort was then analysed for the detection of invasive breast cancer by screening modality, stratified by Volpara breast density groups. The most invasive cancers were detected in women with higher breast density (categories C & D; Table 3). Tomosynthesis identified invasive cancers across all breast density groups, but fewer were detected in women with category D density compared to category C. Ultrasound and MRI detected additional invasive cancers, particularly in women with higher breast density (Table 3). Four women with invasive cancer did not have MRI, which affected the total detections for this modality. Importantly, analysis of invasive cancer detection rates by modality demonstrated substantially higher detection with supplementary imaging. While 2D mammography and tomosynthesis detected invasive cancers at rates of 0.18% and 0.33%, respectively, ultrasound identified invasive disease in 2.1% of the 523 women screened, and MRI detected cancers in 4.7% of the 170 women who underwent MRI (Table 3).

Mammography and tomosynthesis were effective at detecting ductal carcinoma *in situ* (DCIS), likely due to their ability to identify calcifications, which are characteristic of many DCIS cases. However, MRI detected three additional cases of DCIS and one of LCIS that were occult on mammography/tomosynthesis, emphasising its ability to identify non-calcified *in situ* cancer (Table 4). Two women with DCIS detected by mammography/tomosynthesis did not undergo MRI, meaning the additional DCIS detection by MRI occurred in cases where DCIS had not already been identified. These findings reinforce MRI’s complementary role in detecting DCIS, particularly in cases where lesions lack calcifications.

**Table 4:** Analysis of the screening pathway cohort showing the presence of DCIS/LCIS detected by each modality, by TC8 Risk ANZ category or breast density. Note: For the eight women where DCIS/LCIS was detected, six had *in situ* disease in addition to invasive disease, and two had DCIS only.

|  | Mammogram<br>(n=8) | Tomosynthesis<br>(n=8) | MRI<br>(n = 4) | *P*<br>value |
| --- | --- | --- | --- | --- |
| ***In situ* cases detected by TC8 Risk ANZ Category** |  |  |  |  |
| Average | 3 | 3 | 2 |  |
| Intermediate | 1 | 1 | 2 |  |
| High | 0 | 0 | 0 |  |
| ***In situ* cases detected by Breast Density Group** |  |  |  |  |
| A | 0 | 0 | 0 |  |
| B | 3 | 3 | 2 |  |
| C | 1 | 1 | 0 |  |
| D | 0 | 0 | 2 |  |
| **Total by Modality (%)<sup>1</sup>** | 4 (50) | 4 (50) | 4 (100)<sup>2</sup> |  |

<sup>1</sup> Percentage refers to number detected by total screened for each modality.
<sup>2</sup> Four women with DCIS detected by Mammogram/Tomosynthesis did not proceed to have MRI, but MRI detected three additional cases of DCIS and one of LCIS.

Analysis of DCIS detection by breast density showed that DCIS was detected by mammography and tomosynthesis across all breast density groups, except for category A, where no cases were identified (Table 4). The number of DCIS detections was similar between mammography and tomosynthesis, with most cases found in breast density groups B and C. Fewer DCIS cases were detected in women with category D density compared to category C (Table 4). MRI identified additional DCIS cases, with detections in women with categories B, C, and D breast densities.

### Extent of Disease by Screening Modality

To evaluate how each imaging modality contributed to assessing the extent of invasive disease across the full cohort, including both screening and diagnostic pathway cases. Ultrasound identified invasive disease not visualised on either mammography or tomosynthesis in four women.

MRI provided the highest sensitivity and specificity of all the modalities, identifying a larger extent of disease than ultrasound in seven cases, either by increasing the measured lesion size or revealing additional lesions (multifocal disease) not seen on other imaging modalities (Table 5). These results suggest that while ultrasound enhances extent-of-disease assessment compared to tomosynthesis, MRI offers the most accurate assessment for local staging.

**Table 5.**
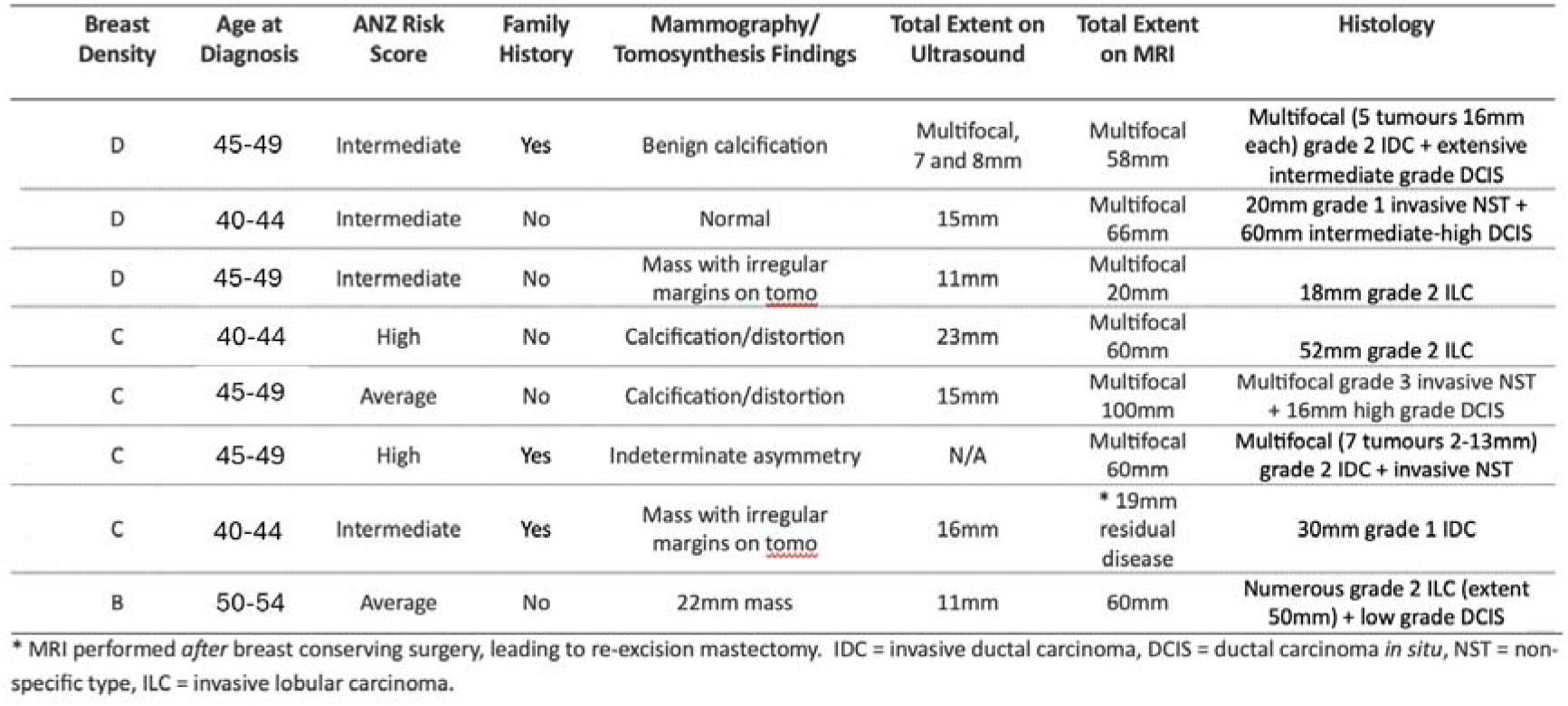
Eight cases where MRI increased the extent of disease detected.

Among the eight women in whom ultrasound and/or MRI identified a greater extent of invasive disease than mammography, seven (87.5%) had dense breasts (Volpara categories C or D), including four with category C and three with category D density (Table 5). Only one case involved a woman with non-dense breasts (category B). The median age at diagnosis was 46 years, and seven of the eight women were under the age of 50, indicating a predominance of younger women and need for MRI assessment in these women to determine total extent of disease prior to surgical management. The TC8 Risk ANZ groups varied for these women, with four women classified as Intermediate risk, two as High, and two as Average risk, suggesting that age and breast density were more consistent markers among these cases than risk score (Table 5). Five women diagnosed with breast cancer had a family history of breast or ovarian cancer (5/19 = 26%), and three of these had a greater extent of disease detected by MRI (3/19 = 16%).

One case demonstrating both breast cancer detection and extent of disease by different screening modalities is shown below. This case, with category D breast density, had invasive breast cancer identified by ultrasound and MRI after being occult on mammography (Figure 2). MRI detected the two invasive cancers seen on ultrasound but also detected a further three invasive cancers and extensive DCIS which were all occult on mammogram and ultrasound (Figure 2). This considerably changed the extent of disease, and surgical management, while correlating well with extent of disease on post surgical histology.

**Figure 2.**
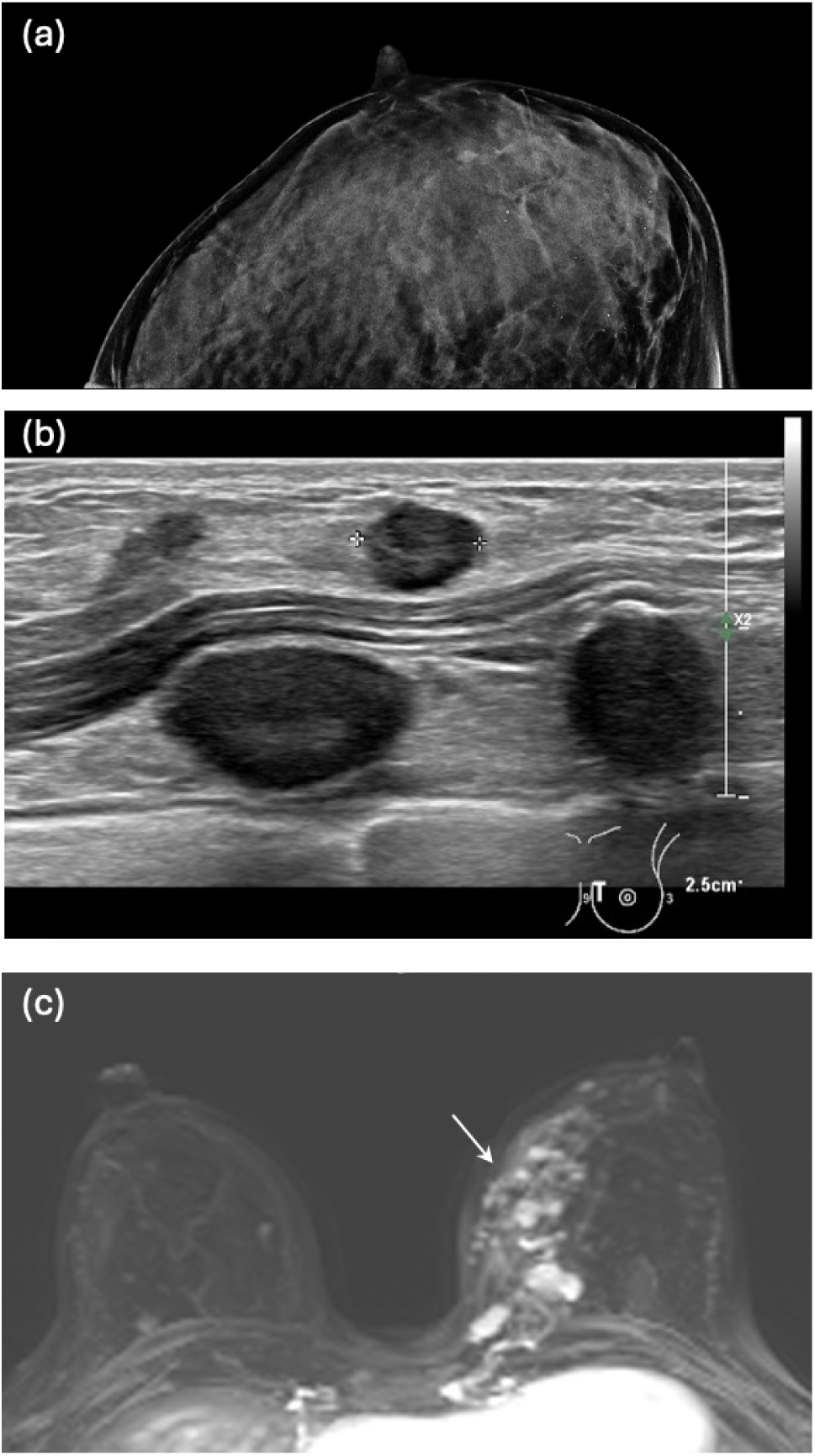
Imaging data from a woman in her 40’s within the screening cohort, classified as ANZ Intermediate Risk. (a) Left breast craniocaudal view on the mammogram showed extremely dense (Volpara “D”) breast with no features suspicious for malignancy. Scattered benign microcalcification was evident. (b) Ultrasound recommended in view of extreme breast density. This detected two irregular masses in the left 9 o’clock position which were mammographically occult-mass 1 measured 8mm maximally, mass 2 measured 7mm maximally. (c) Contrast enhanced breast MRI was performed to assess the extent of disease. This identified multifocal disease (as shown by arrow) from posterior to anterior 1/3 of the left breast with a total of five masses (including the two seen on ultrasound), and segmental non mass enhancement measuring 58mm within the region of mass enhancement in the left breast. Histopathology following mastectomy of the left breast revealed five grade 2 invasive carcinoma of no special type (maximal size 16mm), with extensive intermediate grade DCIS.

## Discussion

This study evaluated the contribution of supplementary breast imaging modalities-digital breast tomosynthesis, ultrasound, and abbreviated MRI, to breast cancer detection and disease assessment in a real-world New Zealand cohort stratified by breast density and clinical risk scores. Our findings indicate that breast density, rather than risk score alone, was the strongest predictor of both mammography-occult disease and benefit from supplementary imaging. Importantly, we also demonstrate the critical role of MRI in accurate local staging.

While clinical risk models such as the TC8 Risk Score are increasingly used to guide personalised screening pathways, their practical utility in determining eligibility for supplementary imaging was limited in our cohort. Detection rates for invasive and *in situ* cancer did not differ substantially across TC8 risk groups, and many cancers occurred in women classified as “Average” risk. In contrast, breast density offered a more actionable stratifier: the majority of cancers missed on 2D mammography occurred in women with dense breasts (Volpara categories C and D), consistent with known masking effects of fibroglandular tissue. ^1 18^

Digital breast tomosynthesis improved detection of invasive disease compared with 2D mammography, confirming its role as a valuable first-line modality across all density categories. ^6^ However, tomosynthesis alone failed to detect over one-quarter of invasive cancers, particularly in women with extremely dense breasts. Most of these were subsequently identified via ultrasound or MRI, underscoring the limitations of non-contrast imaging in this population and highlighting the need for supplementary contrast-enhanced techniques. ^19^

Ultrasound provided significant diagnostic benefit in dense breasts, identifying cancers missed by mammography and offering additional detail on lesion size and focality. However, MRI outperformed all other modalities: in every case where it was performed, MRI detected all cancers previously seen on mammography or ultrasound, and additionally identified further invasive or in situ disease. In some cases, MRI visualised disease 5.5 times larger than the extent seen on ultrasound.

The abbreviated MRI protocol used in this study, combining elements of abbreviated and FAST MRI, was particularly effective in improving both detection and staging accuracy. The ultrafast dynamic TWIST sequence captured early contrast enhancement immediately after thoracic aortic enhancement, before background parenchymal enhancement could obscure lesions. ^14, 15^ This timing improves lesion conspicuity and enhances specificity, especially in dense breasts. The scanning protocol also included axial T2, T1 non-fat saturated, delayed fat-saturated sequences, and MIP reconstructions, while omitting diffusion-weighted imaging to shorten acquisition time to approximately 10 minutes.

These results echo findings from the BRAID trial, ^20^ which demonstrated that MRI significantly improved cancer detection in women with dense breasts, including those with normal mammograms. Likewise, Lawrenson *et al*. ^11^ recently highlighted the potential benefit of more stratified approaches to breast imaging in New Zealand, especially outside of national screening programs. Our findings support MRI not only as a detection tool, but also as the most sensitive modality for local staging, particularly when multifocal or multicentric disease is suspected.

This accurate disease mapping has immediate clinical relevance. MRI’s ability to define lesion extent, chest wall involvement, nipple or skin infiltration, and contralateral disease supports optimal surgical planning and reduces the likelihood of reoperation due to positive margins. Our findings align with Aroney *et al*., ^21^ who reported that preoperative MRI altered surgical management in a substantial proportion of cases. Based on these data, we recommend that all women with dense breasts (Volpara C or D) diagnosed with cancer should undergo MRI to inform preoperative planning. Furthermore, given MRI’s detection of mammographically occult cancers in dense breasts, it may also be appropriate to consider contrast-enhanced screening for women with prior breast-conserving surgery and persistent high breast density.

We also evaluated a revised TC8 threshold (≥ 30%) to align with NICE criteria used in Australasia. ^17^ This adjustment successfully reduced over-triage while preserving alignment with locally used ANZ Risk Scores. However, the revised risk classification still failed to discriminate which women benefited most from additional imaging. These findings support a triage strategy where breast density drives access to supplementary imaging, with risk models playing a secondary role in clinical decision-making.

This study has limitations. It was retrospective, conducted in a single private imaging clinic, and relied on clinical judgment and patient preference for modality selection, which may limit generalisability. Not all women received all modalities, particularly MRI, and longer-term follow-up was not available to assess interval cancers or survival impact.

In conclusion, our data support a density-informed multimodal imaging strategy for breast cancer detection and staging. While tomosynthesis and ultrasound improve diagnostic yield beyond 2D mammography, only contrast MRI provided full visualisation of disease in all imaged cancers. Abbreviated MRI protocols incorporating ultrafast sequences maintain high sensitivity at reduced cost and time, making them viable for widespread use. We recommend that MRI be used in all patients with high breast density and a breast cancer diagnosis to optimise staging and surgical management.

## Data Availability Statement

The datasets generated and/or analysed during the current study are not publicly available due to patient confidentiality but are available from the corresponding author on reasonable request and subject to ethical approval.

## Acknowledgements

We thank the radiologists, radiographers, sonographers, and administrative staff at Mercy Radiology for their contributions to this study. We also acknowledge Breast Cancer Cure, Breast Cancer Foundation NZ and Bayer for funding this work.

